# Effects of the COVID-19 pandemic on self-reported 12-month pneumococcal vaccination series completion rates in Canada: An interrupted time-series analysis

**DOI:** 10.1101/2022.06.21.22276720

**Authors:** Katherine M Atkinson, Blaise Ntacyabukura, Steven Hawken, Lucie Laflamme, Kumanan Wilson

## Abstract

**Background:** Routine childhood vaccination improves health and prevents morbidity and mortality from vaccination preventable diseases. There are indications that the COVID-19 pandemic has negatively impacted vaccination rates globally, but systematic studies on this are still lacking in Canada. This study aims to add knowledge on the effect of the pandemic on pneumococcal vaccination rates of children using self-reported immunization data entered into the CANImmunize digital vaccination tool.

**Methodology:** An interrupted time series analysis was conducted on aggregated monthly enrollment of children on the platform (2016-2021) and their pneumococcal immunization series completion rates (2016-2020). Predicted trends before and after the onset of the COVID-19 related restriction (March 1, 2020) were compared by means of an Autoregressive Integrated Moving Average (ARIMA).

**Results:** Pandemic restrictions were associated with changes in self-reported pneumococcal immunization rates amongst the users of the CANImmunize platform. The monthly enrollment of children on the platform decreased by – 1177.52 records (95% CI: –1865.47, – 489.57), with a continued decrease of 80.84 records each month. Self-reported pneumococcal immunization series completion rates had an immediate increase of 14.57% (95% CI 4.64, 24.51) followed by a decrease of –3.54% each month.

**Conclusion:** The onset of the COVID-19 related restrictions impacted enrollment of children in the CANImmunize digital immunization platform, and an overall decrease in self-reported pneumococcal immunization series completion rates. Our findings support that efforts to increase catch-up immunization campaigns so that children who could not get scheduled immunization during the pandemic are not missed.

## 1. BACKGROUND

Immunizing children against routine vaccine-preventable diseases (VPDs) has been among the most effective interventions in reducing childhood morbidity and mortality and indirectly saving the world a substantial economic loss^1^. Between 2000 and 2020, in developing countries, the use of routine childhood vaccines against VPDs such as measles, pneumonia, severe diarrhea, and meningitis has averted approximately 20 million deaths and saved around US$ 350 billion in cost of illness ^2,3^. This achievement was possible because of the successful mobilization of beneficiaries, excellent planning, and execution of immunization programs^4^.

In March 2020, the outbreak of the highly infectious and deadly Coronavirus disease of 2019 (COVID-19) emerged as an unprecedented challenge^5^. Countries’ governments across the world adopted the World Health Organization (WHO)’s public health and social measures for limiting the spread of the SARS-CoV-2 and mitigating its associated health loss^6,7^. Those measures included face mask-wearing; restrictions on public and private gatherings; cautious opening and closure of schools and businesses; domestic movements as well as public transport restrictions, and stay-at-home orders; and international travel restrictions^7^. Two years into the pandemic, the world has already recorded more than 400 million cases, a death toll surpassing 6 million, and the disruption of many preventive interventions such as the routine immunization programs for children ^5,8^.

Disruptions to the routine childhood vaccines started right after the onset of the pandemic, affecting vaccines’ supply and demand, their cold chain, and delivery^9,10^. Consequently, 23 million children did not receive their scheduled vaccines^11^, the number of doses administered globally fell by 31.3%^12^, and vaccination coverage dropped by approximately 7.7% ^13^. A drop in ordering vaccines was also noted in countries such as the USA, where a 50% reduction happened right after the onset of pandemic-related restrictions^9^.

As observed in many countries, the effect of the COVID-19 restrictions on vaccination rates created an initial shock after the onset of restrictions or delayed recovery of the service after a few months of the pandemic. In some countries such as Canada, such data is still lacking. There is a possibility that the national goal of achieving community protection against all VPDs by 2025 could have been disrupted by the onset of the pandemic-related restrictions^14^, as they caused an approximately 80% drop in in-person doctor visits for children’s services including vaccination during the first wave of infections^15^. The lack of knowledge on the possible effects of timely vaccination and the adoption of immunization tools hinders interventions aiming to recover vaccination uptake and may further increase the risk of VPDs, especially during this period when most restrictions have been eased.

In this study, the impact of the pandemic on immunization efforts in Canada was studied. Changes in children’s enrollment on the pan-Canadian digital vaccination platform, CANImmunize, and self-reported pediatric pneumococcal immunization series completion rates were assessed to describe how the pandemic’s related restrictions affected previously observed trends. The availability of this knowledge provides insights into how the pandemic has affected immunization and will guide interventions that aim to control and prevent VPDs during the current stage of the pandemic, as all provinces are alleviating COVID-19 restrictions.

## 2. Methods

### 2.1. Study design and settings

We conducted a quasi-experimental study to assess the impact of the COVID-19 pandemic restrictions on infant vaccination rates in Canada. COVID-19’s related restrictions, the interrupting variable, were considered a natural event affecting everyone in Canada, while vaccination rates in terms of enrollment of children on the CANImmunize platform and self-reported on-time pneumococcal immunization series completion rates were considered the population-level outcomes. We conducted a single cohort interrupted time series analysis to examine these questions.

### 2.2. Source of data

All data was collected from the CANImmunize database, a pan-Canadian digital vaccination tool established in 2016 for supporting Canadians to manage their own immunization information. Data extraction happened in February 2022, and contained information about when every child’s record was created, the number of pneumococcal doses every child received, and the date each vaccine dose was reported by CANImmunize users (parents and guardians) using their accounts and in accordance with the provincial vaccination protocols. Immunization data in CANImmunize is self-reported, and CANImmunize uses children’s information such as age, sex, and jurisdiction to create a custom immunization schedule for every record^16^.

This study included only records of children whose records that contained at least one dose of any pediatric vaccine reported, and excluded records that had incorrect dates of birth. The data management is described in Figure-1. Records for children born from January 2016 to December 2021 were eligible for enrolment analysis, however, only children born from January 2016 to December 2020 were eligible for the analysis for immunization series completion rate – to include only children that had turned 13 months.

**Figure 1.**
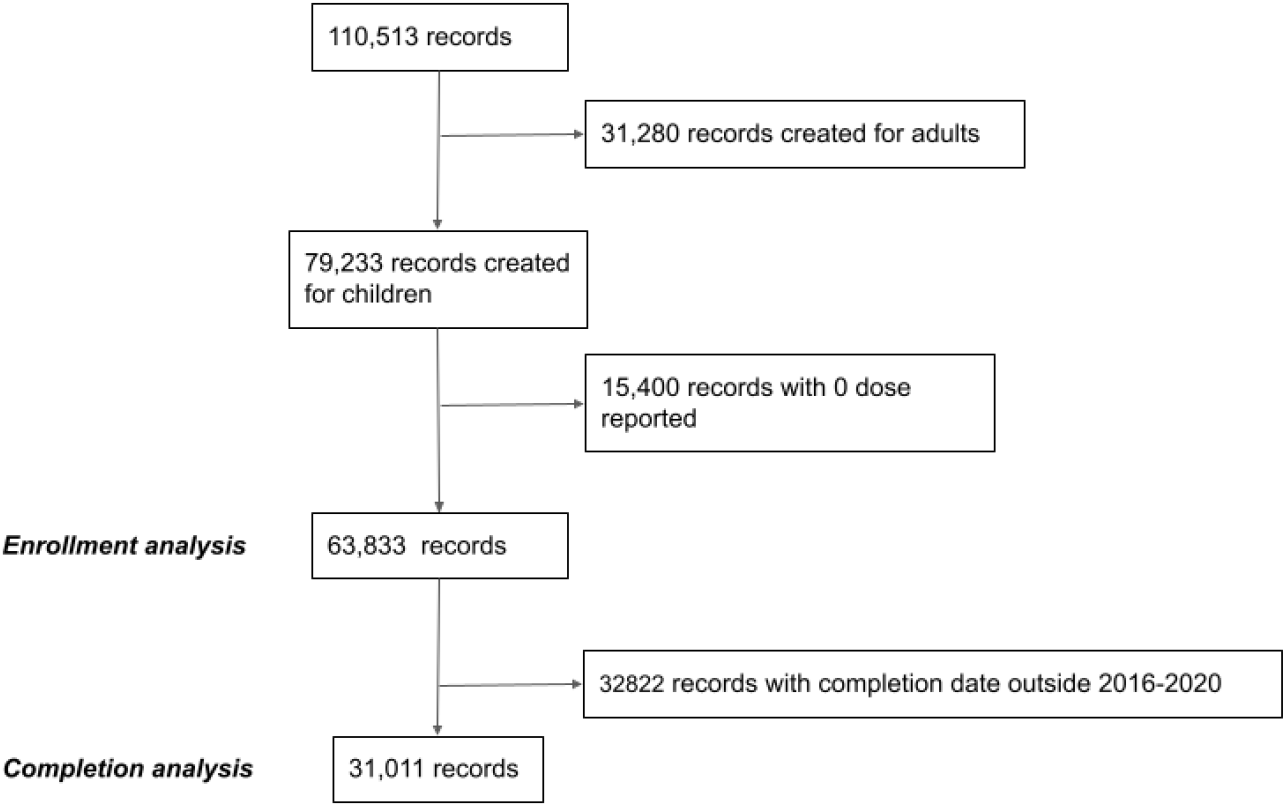
Data management flow illustrating the data cleaning process to obtain aggregated monthly counts of enrolled children and monthly completion rates for pneumococcal vaccine

### 2.3. Statistical analysis

Two outcomes were assessed; Enrollment of children on the CANImmunize platform, which was assessed as the number of records created on CANImmunize; and the on-time completion rate of self-reported immunization series for pneumococcal vaccine. The number of new records was counted monthly using the record’s unique identifiers on CANImmunize. Ontime completion of immunization series for pneumococcal vaccine was defined as reception of three doses of pneumococcal vaccine before the child turns 13 months of age^22^. It was calculated using the child’s date of birth, the date the child turned 13 months, and the number of doses of pneumococcal vaccines every child received in that period.

The count of children that were born every month during the observation period served as the denominator, while the count of children born in the same month and received all doses of pneumococcal vaccine before they turned 13 months served as the numerator to provide the monthly completion rate in percentage.

A total of 52 (from September 2017 to December 2021) and 60 (from January 2016 to December 2020) monthly time series were generated for enrollment and immunization series completion rates, respectively.

We hypothesized that the onset of COVID-19 related restrictions in Canada would impact both outcomes in a step-change format, an assumption based on the interim reports from the World Health Organization (WHO) on the impact of the pandemic on universal immunization coverage^8^. The point estimate and the confidence interval for the impact of the pandemic on both outcomes were assessed using Autoregressive Integrated Moving Average (ARIMA) models.

#### Model identification

After examining the trend and patterns of both outcome’s time series(see Figure 2), the ARIMA model was used to account for autocorrelation and seasonality bias. The Dickey-Fuller test was used to confirm the series’ stationarity^17^ prior to fitting an ARIMA model. We used an automated algorithm, specifically *auto*.*arima()* in the *forecast* package for R, to identify the ARIMA model terms based on minimizing the information criteria (AIC, BIC). The statistical R version 4.1.3 was used for all analyses and both the P_value and the confidence interval were reported. The *P*_value less than 0.05 was considered statistically significant.

**Figure 2.**
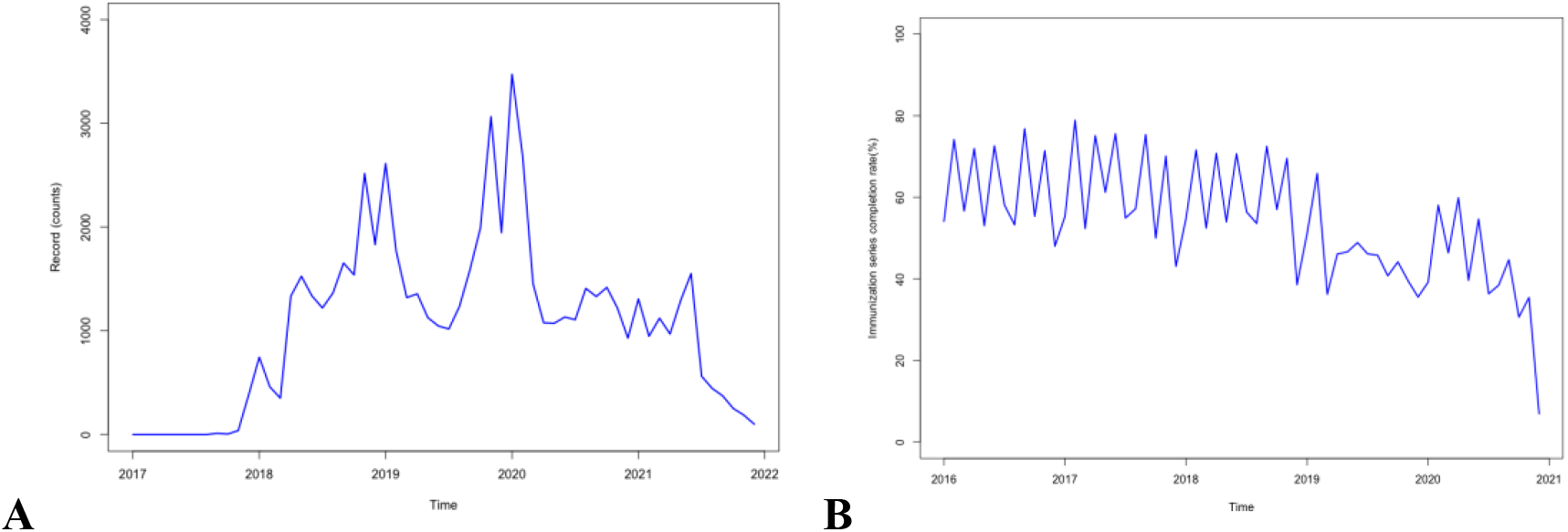
(A) shows the time series of monthly enrollment of children on CANImmunize from January 2016 to December 2021. (B) shows the time series for pneumococcal vaccine’s completion rates in the period from 2016 to 2020.

**Figure 3.**
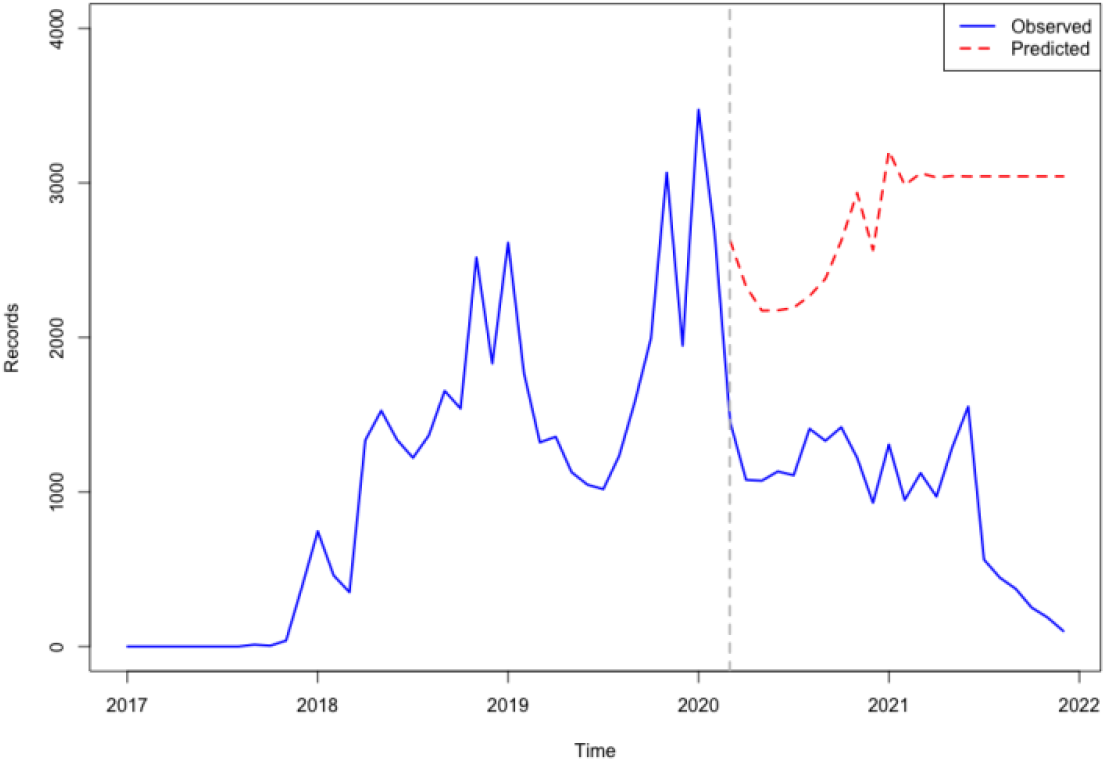
shows both the observed and predicted children’s enrollment on CANImmunize in the period from 2016 to 2021. (The red-dash line represents the cut-off, the onset of Covid-19 related restrictions in Canada^19^).

**Figure 4.**
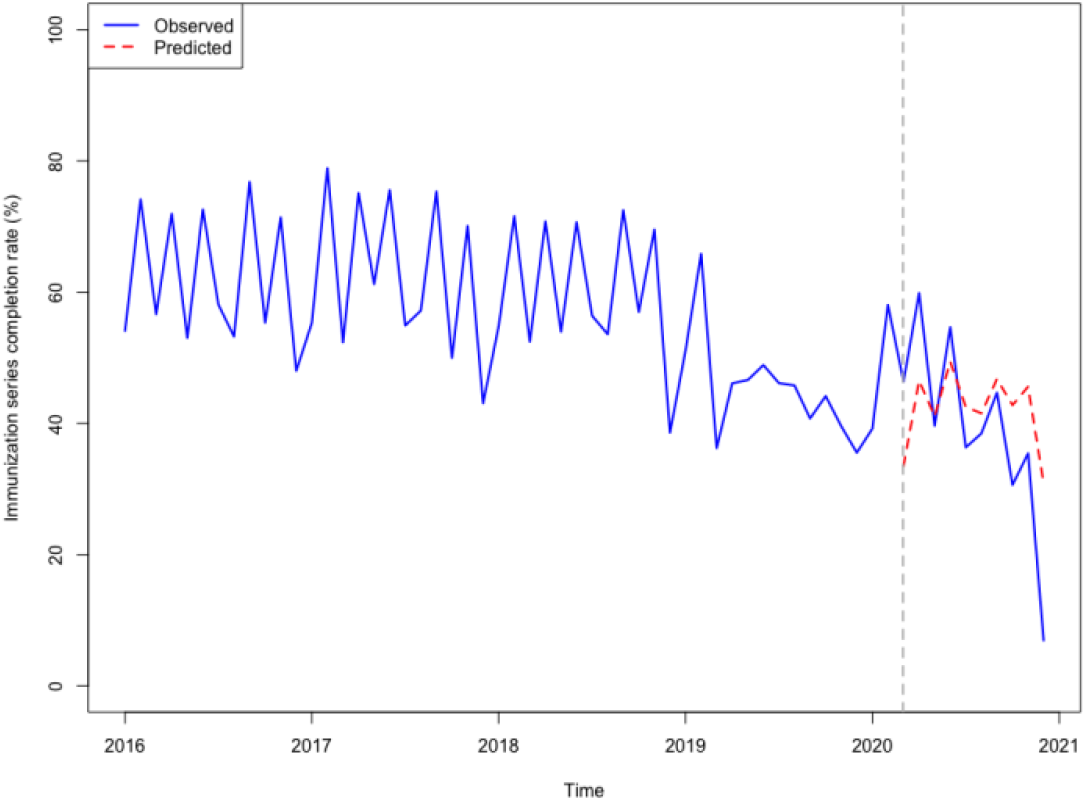
shows the predicted completion rates compared to observed rates from 2016 to 2020. The red-dash line represents the cut-off, the onset of Covid-19 related restrictions in Canada^19^

## 3. Ethical considerations

Data collection and handling were conducted according to the CANImmunize privacy policy^18^. Additionally, ethical approval to conduct this research has been obtained from the Ottawa Health Sciences Research Ethics Board (OHSN REB) prior to the start of all research activities.

## 4. RESULTS

### 4.1. Overview of our data

A total of 63833 pediatric records were created in CANImmunize from 2016 to 2021have met inclusion criteria. The highest enrollment of children occurred in January 2019 with 3374 records as *Figure 2*(A) shows. The time series for trend for enrollment displayed seasonality features including a spike in the number of enrolled children towards the end of each year. However, there was a decrease in counts of enrolling children observed after the onset of COVID-19 restrictions, and the tall spikes observed in the pre-restriction period have not been repeated. Additionally, the post restriction trend did not recover to reach the pre-restriction level; further decreasing until the end of the study period. The trend for pneumococcal immunization series completion rates has been decreasing since the end of 2018, and the recovery that happened in the early months of 2020 was not sustained.

### 4.2. ARIMA model

#### (1) Enrollment series

The estimated step change was –1177.52 records (95% CI: –1865.47, – 489.57) while the estimated change in slope was -80.84 records per month (95% CI - 227.03, 65.34). Figure 5 shows the values predicted by our ARIMA model in the absence of the intervention (counterfactual) compared with the observed values. This means that restrictions that were imposed to curb the impact of COVID-19 in March 2020 were associated with an abrupt and continued decline in enrollment by 1177.52, with a further decrease of 80.84 records each month during the study period. The p-value for the Ljung-Box test for white noise is 0.73 at 24 lags.

#### (2) Pneumococcal series completion rates

The estimated step change was 14.57% (95% CI 4.64, 24.51)

while the estimated change in slope was –3.54% per month (95% CI -5.31, –1.76). Figure 5 shows the values predicted by our ARIMA model in the absence of the intervention (counterfactual) compared with the observed values. This means that restrictions that followed the onset of COVID-19 restrictions in March 2020 were associated with an increase in self-reported completion of scheduled penumococcal immunization series by 14.57%, followed by ar decrease of 3.54% each month during the study period. The p-value for the Ljung-Box test for white noise is 0.98 at 24 lags.

## 5. DISCUSSION

The onset of COVID-19 in March 2020 in Canada was associated with changes in immunization rates among users of the CANImmunize platform. The monthly enrollment of children on the platform has dropped by – 1177.52 records (95% CI: –1865.47, – 489.57), with a continued drop of 80.84 records each month. Pneumococcalimmunization series completion had an initial increase of 14.57% (95% CI 4.64, 24.51) and a sustained decrease of –3.54% each month, producing an estimated net effect of -20.83%.

These findings are consistent with results from studies emerging on the impact of the COVID-19 pandemic on routine childhood vaccines globally. In North America, studies in the United States of America, and the Dominican Republic have reported a decline of 5-18%, and 10% in vaccination coverage, respectively^20–24^. In South America, studies in Columbia and Brazil have reported a drop in vaccination coverage of 14.4-20% and 10 to 20%, respectively^25^. European countries, in general, saw a 1% decrease in vaccination coverage, while the African region saw a 2-18% decrease^26,27^. Middle eastern countries such as Jordan have reported a decrease in vaccination coverage of 11.1%^28^, while Lebanon reported a national level decrease of 31% in the utilization of immunization services during the pandemic^29^.

To our knowledge, there are no studies that assessed the impact of the pandemic-related restrictions on population engagement in digital tools for vaccination such as CANImmunize and the impact of the pandemic on the pneumococcal vaccination in Canada. A key strength of our study is that we used a large dataset, observed over a long period. It has allowed us to compare and understand changes in the trend of each outcome. Additionally, this research was the first to detect the effect of the COVID-19 pandemic on children’s immunization in Canada, and it will serve as hypothesis-generating while other nationwide evaluations are conducted.

Thisur study had some limitations. Outcomes were assessed in a population that is actively utilizing a free, digital tool for managing immunization records, who may not be representative of the general population. Disparities in immunization rates according to geographical and social-economic status were not assessed. Our study was also a single-group interrupted time series analysis, lacking a parallel comparison group. Furthermore, all data on vaccination was self-reported However, vaccinations reported by parents are used by some provinces as the official source of data. Parents are required to register such information to public health for school entry under the *Immunization of School Pupils Act* in Ontario^30^.

## 6. CONCLUSION

The COVID-19 pandemic has negatively affected enrollment in the CANImmunize app immediately and the whole time under pandemic restrictions. Pneumococcal immunization series completion rates had an initial increase but an overall estimated net effect of -20.83%. Further research is needed to assess the impact of the COVID-19 pandemic on pediatric vaccination rates in Canada. There is a strong need for the development and implementation of catch-up intervention programs to mitigate the impact of the pandemic on the 2025 Public Health Agency of Canada goals of achieving community protection against VPDs.

## Supporting information

Supplemental images illustrating outputs of the statistical analysis

## Data Availability

All data produced are available online at: Ntacyabukura, Blaise (2022), Children enrolment on CANImmunize and the pneumococcal vaccine's completion rate dataset, Mendeley Data, V1, DOI: 10.17632/bxspw2d2wx.1

## 9. ACKNOWLEDGMENT

We thank the team of supervisors from Karolinska Institutet for their guidance during the research proposal writing and data analysis.

