## Supplementary figures and images for "Effects of the COVID-19 pandemic on self-reported 12-month pneumococcal vaccination series completion rates in Canada: An interrupted time-series analysis"

### Supplemental images illustrating outputs of the statistical analysis

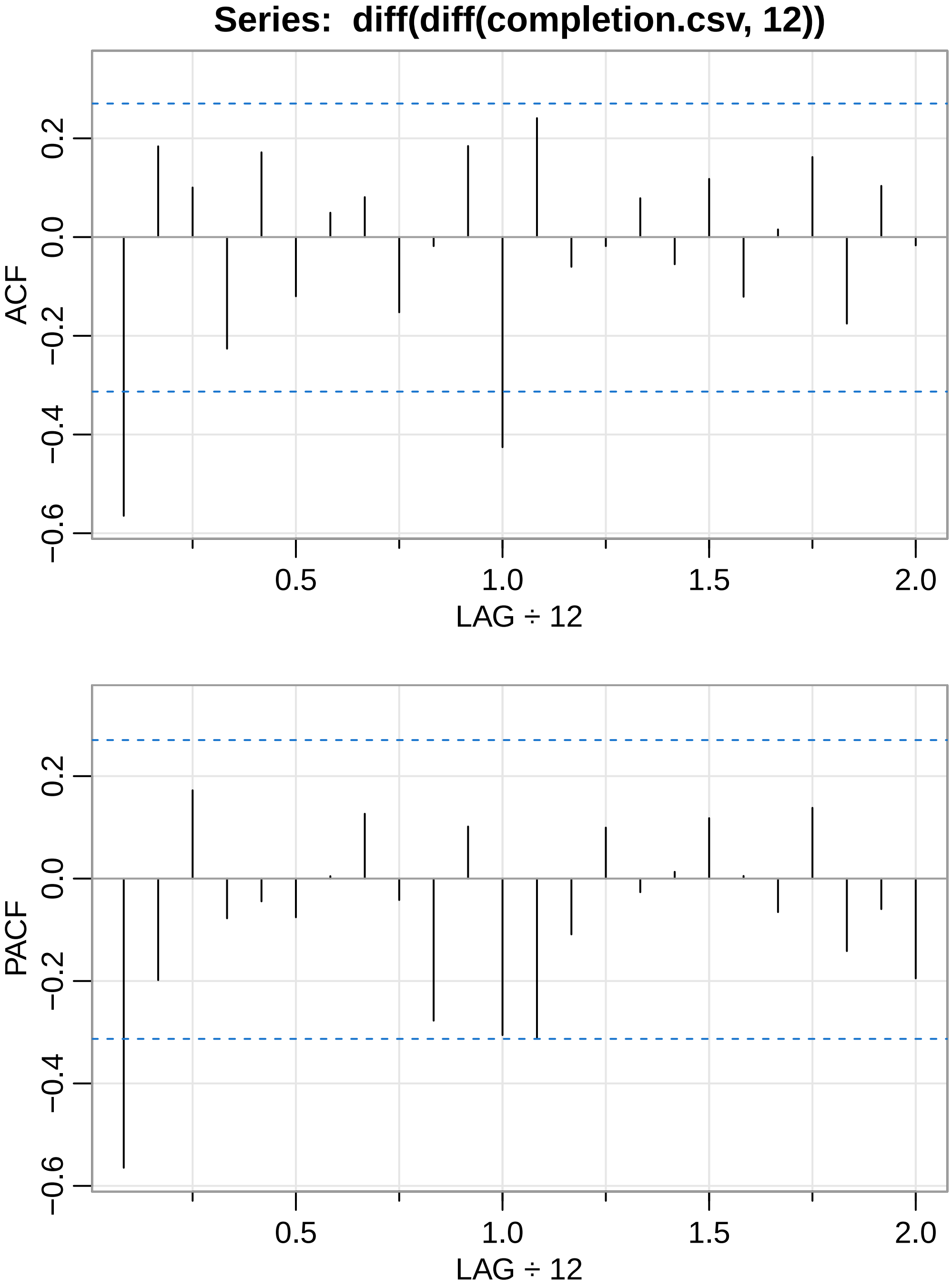

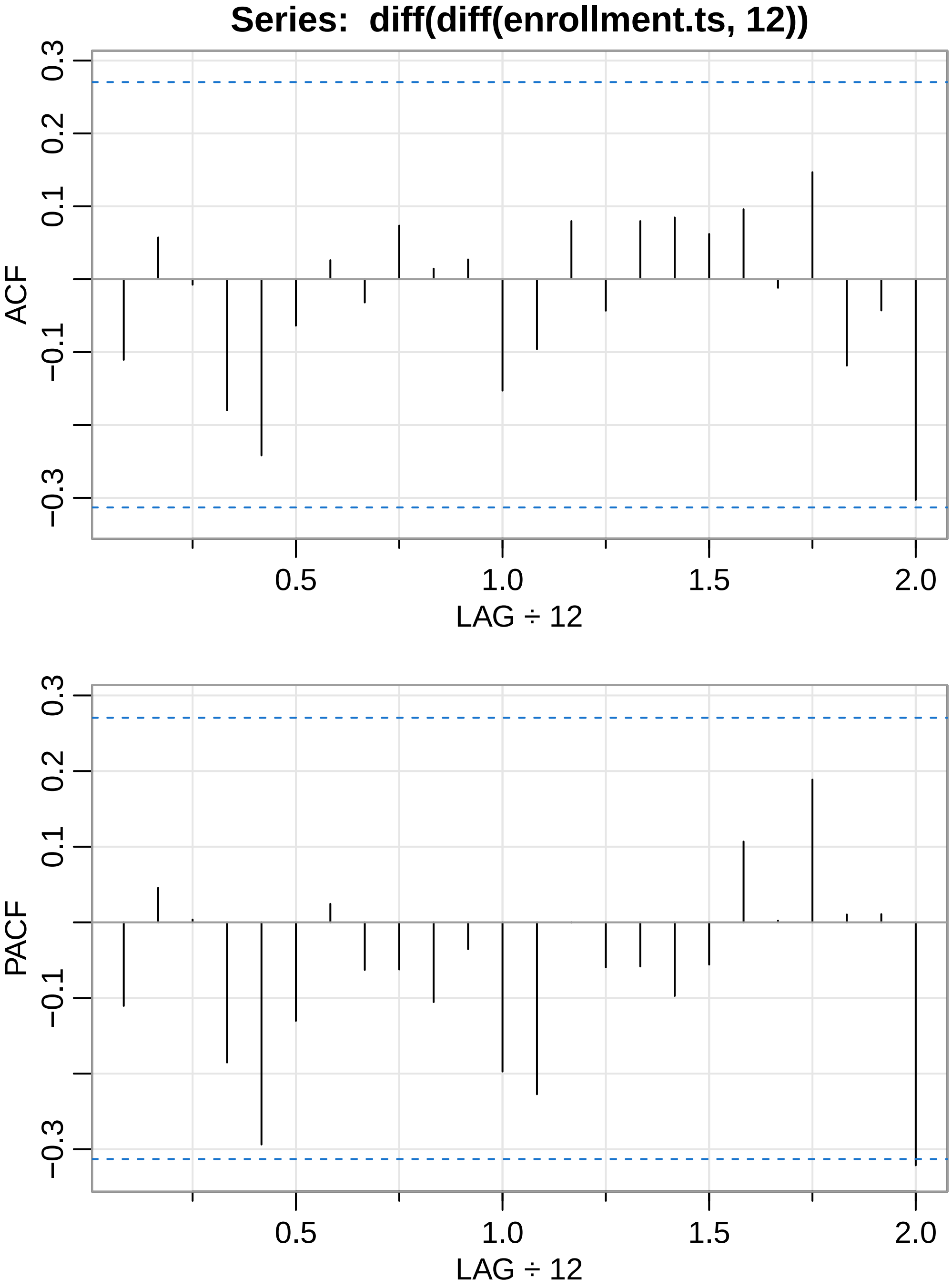

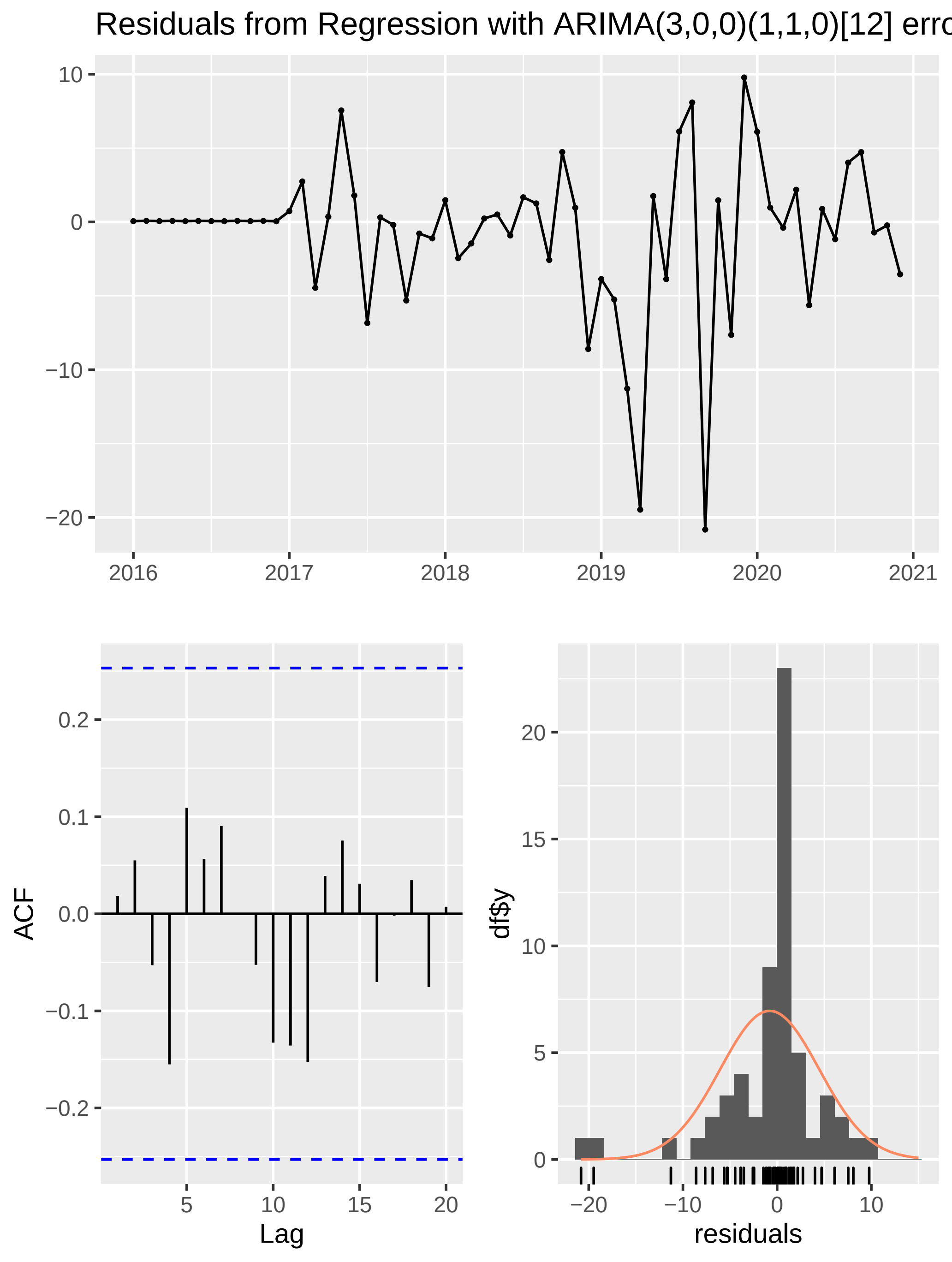

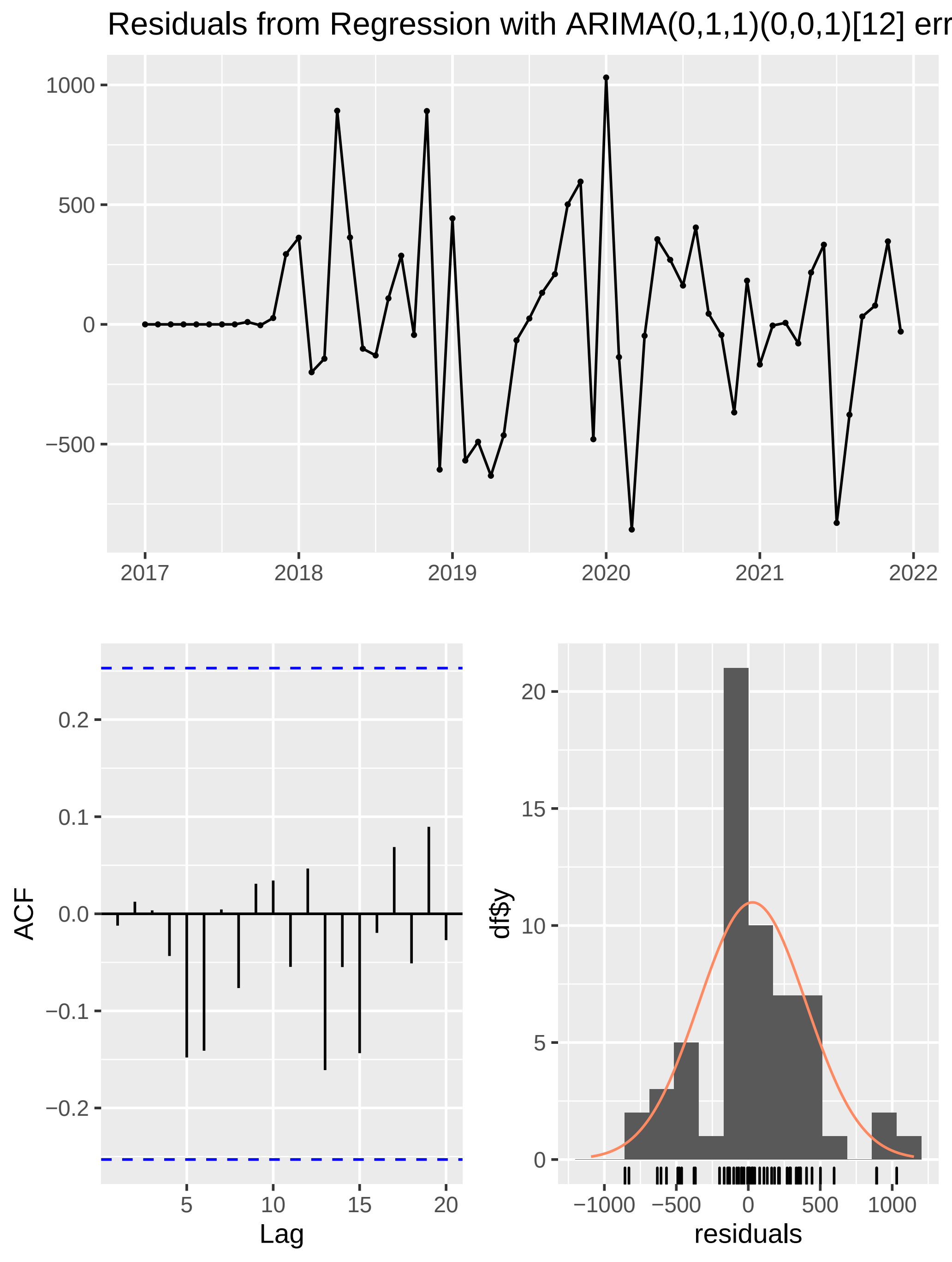
